# Tracking Respiratory Syncytial Virus dynamics in wastewater during the 2024-2025 season in Switzerland

**DOI:** 10.64898/2026.05.14.26352723

**Authors:** Auguste Rimaite, Jolinda de Korne-Elenbaas, Adrian Lison, Tanja Stadler, Timothy R. Julian, Niko Beerenwinkel

## Abstract

Respiratory Syncytial Virus (RSV) is responsible for a substantial health burden worldwide, particularly among children and older adults. In 2023, novel immunoprophylactic interventions for RSV were approved, underscoring the need to monitor circulating RSV lineages and detect mutations that could compromise intervention effectiveness. Here, we implemented wastewater-based genomic RSV surveillance by integrating digital PCR and amplicon-based sequencing within Switzerland’s national wastewater monitoring program. We tracked RSV subtypes and individual mutations across the 2024-2025 peak season in six Swiss cities. RSV-A and RSV-B co-circulated nationwide, and both exhibited similar epidemiological dynamics estimated from their subtype-specific effective reproduction numbers. No previously reported F protein mutations relevant to prophylaxis efficacy were identified. Genetic diversity analysis of wastewater-derived sequences reflected patterns previously reported in clinical data, with higher diversity in RSV-A than RSV-B and greater variability in the G compared to the F gene. These findings demonstrate the potential of wastewater-based RSV surveillance for monitoring RSV dynamics and diversity and establish a national baseline for RSV evolution during the first season following vaccine implementation in Switzerland.

## Introduction

Respiratory Syncytial Virus (RSV) is one of the most common respiratory pathogens, causing acute lower respiratory tract infections. While typically causing mild cold-like symptoms, RSV infections can develop into severe respiratory disease in vulnerable groups like young children and older adults^1,2^. The associated need for hospitalization burdens healthcare systems, particularly during winter months when RSV circulation peaks alongside other respiratory viruses^3^. Recently approved RSV vaccines and monoclonal antibodies effectively protect infants against severe disease in their first RSV season, thereby reducing hospitalization rates^4,5^.

RSV consists of two major antigenic subgroups, RSV-A and RSV-B, differentiated based on the G glycoprotein, a major surface protein that, together with the fusion (F) glycoprotein, is responsible for viral infectivity^6,7^. The F glycoprotein is highly conserved and therefore the primary target of immunoprophylaxis. RSV vaccines contain a stabilized prefusion form of the F protein (preF), offering neutralizing activity against both RSV subtypes^8–10^. Similarly, the long-acting monoclonal antibody nirsevimab targets a highly conserved region of the F protein with demonstrated efficacy against both subtypes^11,12^. The widespread use of these interventions introduces new selective pressures on circulating RSV strains, potentially impacting subtype dynamics, or selecting for escape mutations^13^. This highlights the need for genomic surveillance of circulating strains to ensure continued prophylaxis effectiveness.

RSV cases and genomic surveillance are used to guide and evaluate intervention strategies. In many countries, RSV surveillance systems are focused on vulnerable populations in hospitals and older adult care facilities. In Switzerland, RSV monitoring is based on voluntary reporting by clinicians through the Sentinella system and RSV EpiCH network, leaving gaps in knowledge on RSV circulation within communities^14,15^. Wastewater-based surveillance is a powerful complementary surveillance tool as total RSV loads in wastewater mirror trends in positivity rates^16,17^ and reported clinical cases^18,19^, and subtype-specific assays allow tracking of RSV-A and RSV-B individually^20^. The subtypes are thought to alternate in predominance every year; however, their dynamics are not regularly monitored. With the introduction of new RSV prophylaxis, the need for genomic surveillance increased to capture population-level infection trends and potential resistance mutations, ensuring informed public health decision-making and continued effectiveness of prevention strategies. Wastewater-based genomic surveillance of RSV has proven effective in population-level monitoring of circulating lineages and mutations at prophylaxis-relevant sites^17,21^.

Here, we implemented wastewater-based genomic surveillance of RSV, integrating digital PCR (dPCR) and amplicon-based sequencing within the Swiss national wastewater monitoring program during the 2024-2025 RSV season. Wastewater sequencing data allowed population-level tracking of RSV subtypes and mutations at clinically relevant sites in the F gene, as well as characterization of genetic diversity among subtypes and genes. Altogether, this study provides a basis to explore RSV evolution during the first season following the implementation of novel vaccines in Switzerland in autumn 2024^22^.

## Methods

### Wastewater sampling and processing

Twenty-four-hour composite samples of raw influent wastewater were collected from wastewater treatment plants in six cities across Switzerland: Basel, Chur, Geneva, Lugano, Laupen, and Zurich, as part of Switzerland’s wastewater-based respiratory virus surveillance program^23^. Total nucleic acids were extracted and concentrated from 40 mL raw wastewater sample into 80 μL of eluate using the Wizard® Enviro Total Nucleic Acid Extraction Kit (Promega Corporation, Madison, USA) as previously described^18^. To reduce PCR inhibition, the extracts were purified using OneStep PCR Inhibitor Removal columns (Zymo Research, Irvine, USA) and diluted three-fold. During the RSV peak season (8 Nov 2024, to 30 Apr 2025), three extracts per week were selected for sequencing from each location in 2024 and two per week from each location in 2025.

### Quantification of pan-RSV and subtypes using digital PCR

Pan-RSV concentrations were quantified in the wastewater extracts by targeting a conserved region of the RSV N-gene as part of a sixplex dPCR assay, as described previously^18^. The RSV subtypes RSV-A and RSV-B were quantified using a duplex dPCR assay targeting a subtype-discriminative region of the N-gene, as described previously^21^. A sample was considered to have passed quality control if the number of analyzable droplets generated during dPCR was above the threshold (pan-RSV assay: 15,000 droplets before 25 March 2025, and 12,000 droplets after 25 March 2025; RSV subtyping assay: 12,000 droplets throughout the season). Concentrations were normalized by total daily flow values and catchment population size to obtain loads in genome copies per day per person in the catchment. Pan-RSV concentrations were measured in four samples per week, and RSV was subtyped in two samples per week throughout the season.

### R_t_ estimation

Measured subtype concentrations were used to estimate the time-varying effective reproduction number R_t_ using a Bayesian semi-mechanistic wastewater model described in Lison et al. and implemented in the R package EpiSewer^24^. The generation time distribution of RSV was modeled using a shifted Gamma distribution (mean: 7.5 days, sd: 2.1 days) based on estimates from Vink et al.^25^. The shedding load distribution was modeled using a Gamma distribution with uncertain parameters (mean: 4.4-9.2 days, coefficient of variation: 0.25-0.45) based on nasal swab viral loads measured by Otomaru et al^26^. The total detectable load per infection was calibrated to catchment-specific pan-RSV prevalence numbers estimated using publicly available Sentinella data in Switzerland (Table S1). The same assumptions for all values (generation time distribution, shedding load distribution, and total detectable load per infection) were used when estimating R_t_ for RSV-A and RSV-B. R_t_ was smoothed using a Gaussian process prior as described in Lison et al^24^. The concentration measurements were modeled using a dPCR-specific likelihood that accounts for the volume and number of partitions of the assay^27^. Posterior samples of Rt were obtained via Markov Chain Monte Carlo sampling using the No-U-Turn Sampler (NUTS) in cmdstan version 2.34.1^28^ with 4 chains and 1000 warm-up and 1000 sampling iterations each. Diagnostic checks showed good E-BFMI values (>0.3), few divergent transitions (<1.5%), and sufficient mean R-hat statistics (<1.035) for all subtypes and locations. R_t_ estimates were summarized using the posterior median and 50% and 95% credible intervals (CrIs). Correlation between subtype-specific median R_t_ estimates was assessed using the Pearson correlation coefficient.

### RSV sequencing

RSV-A and RSV-B was amplified and sequenced from the wastewater extracts using a subtype-specific amplicon-based sequencing method, as previously described^21^. Until 19 Nov 2024, only the predominant subtype at each location was sequenced under the assumption that sequencing performance is higher when using only one primer pool during periods of low concentrations. From this date onward, both subtypes were sequenced (Figure S1). Sequencing was performed weekly in 2024 (when three extracts per week per site were sequenced) but reduced to every other week in 2025 (when two extracts per week per site were sequenced). Amplicons of RSV-A, RSV-B, Influenza A, and SARS-CoV-2 were pooled at equal volume ratios in all sequencing runs. The 400 base pair amplicons were sequenced on the AVITI (Element Biosciences) sequencing system, yielding paired-end reads of length 250 bp.

### Mutation frequencies

Raw sequencing data were processed using V-pipe^29^ version 3.0.0.pre1, as previously described^21^. Genome coverage was defined as the proportion of reference genome positions to which at least 10 sequencing reads align. Mutations relative to reference genomes EPI_ISL_412866 (RSV-A) and EPI_ISL_1653999 (RSV-B) were called using LoFreq version 2.1.3^30^, and mutation frequencies were calculated as the number of reads supporting the mutation divided by the total read depth at that position. Positions covered by fewer than 10 reads were treated as missing values^31,32^. In the heatmaps, only non-synonymous mutations observed in at least two wastewater samples at frequencies above 0.02 were visualized. Nucleotide changes were translated into amino acid substitutions using a modified version of vcf-annotator^33^. Clinical sequences were obtained from the Pathoplexus database^34^, including sequences from Switzerland (RSV-A n=34, RSV-B n=42; accession date: 12 Oct 2025) and European countries (RSV-A n=347, RSV-B n=431, accession date: 13 Oct 2025), collected between October 2024 and May 2025. F-gene substitution frequencies in clinical samples were calculated as the proportion of sequences with the mutation relative to the total number of sequences with a valid (non-gap and non-ambiguous) nucleotide at that position.

### Comparing genetic diversity between subtypes and genes

Genetic diversity was quantified using common metrics^35,36^, including mutation richness and nucleotide diversity. Because diversity metrics are sensitive to sequencing depth^36^, diversity estimates included only samples with ≥50% genome or gene coverage at ≥ 10 reads, and only positions covered by ≥10 reads were considered. Nucleotide diversity was computed as 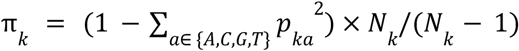 where *pka* is the frequency of nucleotide *a* at site *k*, and *N*_*k*_ is number of reads covering position *k*. Mutation richness was computed as the total number of mutations per kilobase of the covered genome. Mean π values were computed by averaging the positional diversity values defined above over all covered sites in the genes or whole genome.

Differences in diversity between subtypes and genes were assessed using generalized linear mixed effects models (GLMM) with a Gamma distribution and logarithmic link function, implemented in the R package glmmTMB (v1.1.14). The Gamma family was chosen to account for the right-skewed, positive continuous distribution of diversity values. To account for the effect of sequencing depth on observable genetic diversity, linear and quadratic terms of the centered and scaled log-transformed mean positional coverage were included as fixed effects. All fitted models comparing the diversity across subtypes and genes are summarized in Table S2. Mutation richness was not compared between subtypes since this reference-dependent metric reflects cumulative divergence from the reference. A comparison would therefore be confounded by the differences in divergence time from the reference genome. Estimated marginal means and pairwise contrasts were obtained using the emmeans R package (v2.0.1) and back-transformed to the original scale (Tables S3, S6). P-values were adjusted for multiple comparisons using the Benjamini-Hochberg method.

## Results

### Spatial and temporal variation in RSV subtypes across cities

We measured total RSV and subtype-specific concentrations in 289 selected wastewater samples. The median ratio of pan-RSV to combined RSV-A and RSV-B concentrations was 0.97 (range: 0-4.61) (Figure S2A), varying from 0.67 to 1.12 across locations (Figure S2B), indicating that they generally aligned well. RSV loads were highest in Zurich and Geneva and lowest in Laupen (Figure 1). On average throughout the season, RSV-A dominated in Zurich (median proportion 0.70, IQR 0.53–0.77) and Lugano (median 0.70, IQR 0.53–0.85), and was slightly more abundant than RSV-B in Chur (median 0.66, IQR 0.50–0.83) and Laupen (median 0.58, IQR 0.46–0.90). In Basel, the subtypes circulated in roughly equal proportions (median RSV-A 0.49, IQR 0.28–0.63), whereas RSV-B was slightly more prevalent in Geneva (median 0.64, IQR 0.48–0.74) (Table S4). Lineage patterns were broadly consistent across sites, with A.D.1 and B.D.E.1 being predominant throughout the season (Figures S3-S6).

**Figure 1.**
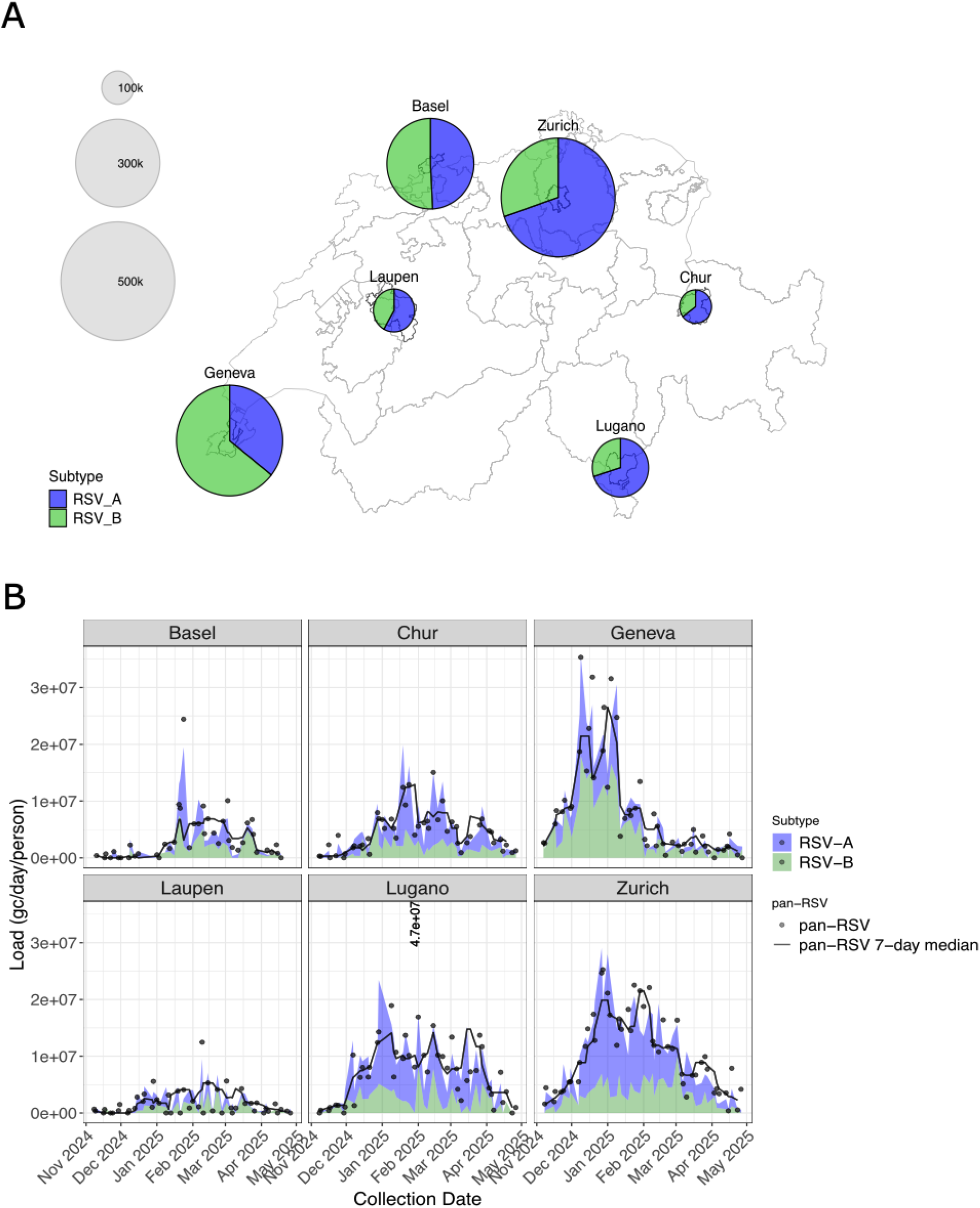
Spatial and temporal dynamics of RSV subtypes across six locations in Switzerland during the 2024-2025 season. (A) Map of Switzerland indicating the locations of the six wastewater treatment plants sampled in this study. Disc sizes are scaled by the population size of each catchment area, and colors represent RSV subtypes (blue for RSV-A and green for RSV-B). Colored fractions indicate the median subtype proportions estimated from subtype-specific concentrations over the study period. (B) Loads of pan-RSV, RSV-A, and RSV-B over time across the six locations. Concentrations were measured using two different dPCR assays targeting either a conserved region of the RSV N gene (part of a sixplex respiratory virus dPCR assay) or a subtype-discriminative region of the RSV N gene. Concentrations were normalized by daily flow and population size of the catchment area to obtain loads in genome copies per day per person. The line represents the pan-RSV 7-day rolling median. The loads of RSV-A and RSV-B are stacked. For visualization purposes, the highest load measured in Lugano (10 Feb, 2025) is not plotted but instead indicated as a label.

### Subtype-specific R_t_ estimates

Estimates of the effective reproduction number R_t_ suggest highly similar transmission dynamics for RSV-A and RSV-B (Figure 2). Across all cities, we found R_t_ to be significantly above 1 at the start of the measurement period for both subtypes. On 1 Nov 2024, median R_t_ estimates ranged from 1.21 to 1.71 for RSV-A and from 1.30 to 1.58 for RSV-B across locations. Moreover, R_t_ trajectories over time were strongly synchronized between the subtypes. Specifically, while the estimated peak of RSV-A infections (i.e., first date for which 50% CrI of Rt < 1) varied significantly across locations (e.g., 19 Dec 2024, in Geneva; 1 Feb 2025, in Basel), the peak of RSV-B infections occurred within 3 weeks of the RSV-A peak (Chur: 15 days earlier, Lugano: 6 days earlier, Zurich: same day, Basel and Geneva: 1 day later, Laupen: 20 days later). Across the measured time period, median R_t_ trajectories of RSV-A and RSV-B for each city were significantly correlated, with a Pearson correlation coefficient ranging from 0.88 to 0.99 (p < 2.9 × 10^-60^ for all locations).

**Figure 2.**
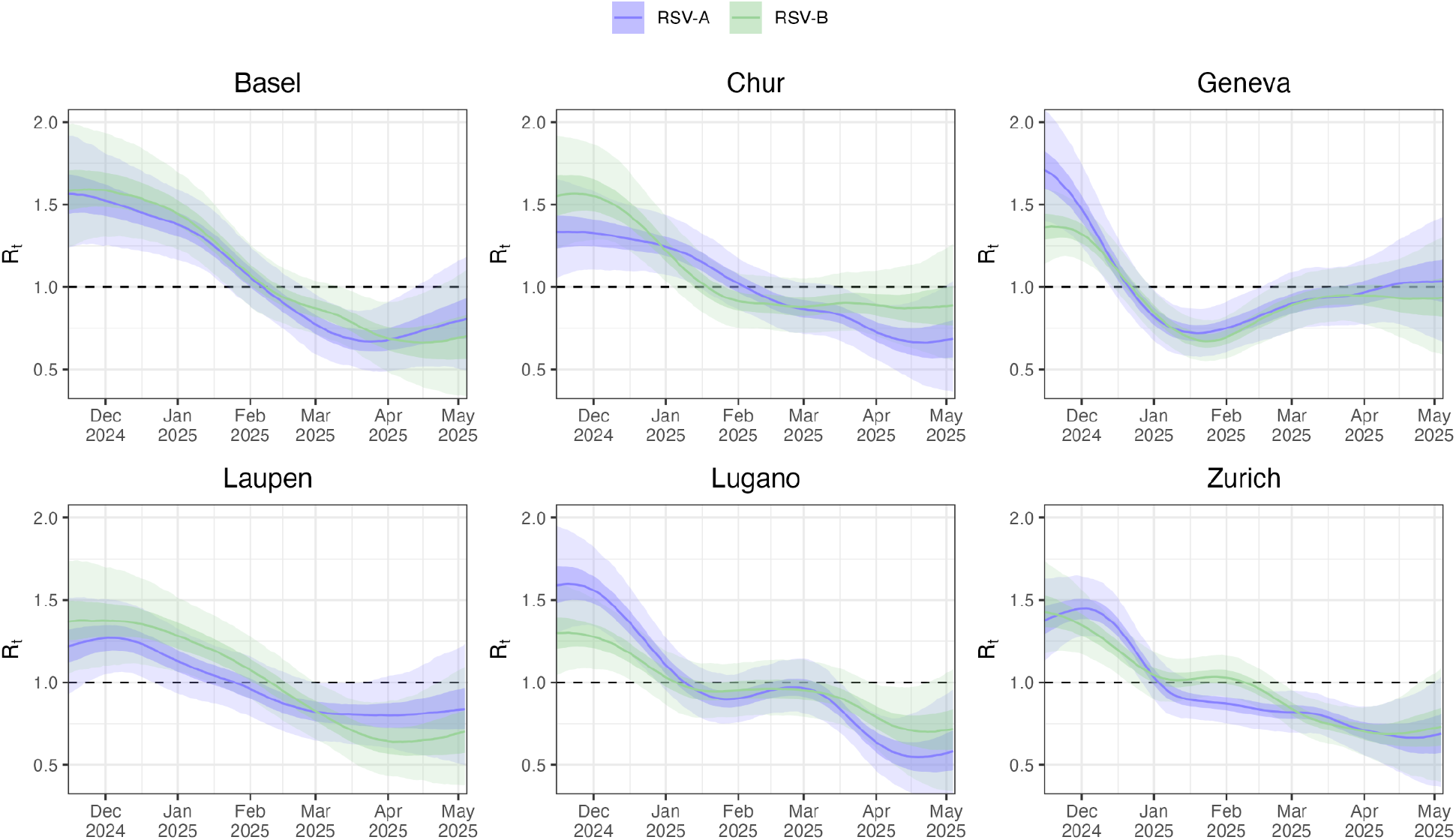
Subtype-specific estimates of the effective reproduction number R_t_. R_t_ estimates for the six Swiss cities between Nov 2024 and May 2025 were obtained from subtype-specific concentration measurements and daily flow data using a Bayesian wastewater model implemented in the R package EpiSewer. Lines show the posterior median; light and dark shaded bands show the 50% and 95% credible intervals of R_t_.

### Wastewater-based RSV sequencing and genome coverage>

We sequenced a total of 328 wastewater samples, covering the RSV season from November 2024 to May 2025, with 48 sequenced for RSV-A, 15 for RSV-B, and 265 for both subtypes (Figure S1). For samples with non-zero subtype concentrations, the median genome coverage (≥10 reads) was 0.16 for RSV-A (range: 0 – 0.89) and 0.17 for RSV-B (range: 0 – 0.91), increasing to 0.34 for RSV-A (range: 0 – 0.89) and 0.31 for RSV-B (range: 0 – 0.91), for samples above the subtyping dPCR assay’s detection limit of six genome copies per mL wastewater^18^. Full genome coverage was strongly correlated with measured subtype concentrations across all locations (RSV-A: ρ = 0.65; RSV-B: ρ = 0.53; p < 2.2 × 10^−16^), as was F-gene coverage (RSV-A: ρ = 0.57, p < 2.2 × 10^−16^; RSV-B: ρ = 0.42, p = 4.88 × 10^−12^) (Figure S7).

### Detection of amino acid substitutions in the prophylaxis-targeted F gene of RSV-A

We compared the frequencies of F-gene amino acid substitutions observed in wastewater throughout the season to their corresponding frequencies in clinical sequences. For RSV-A, we detected three substitutions in wastewater that were present in clinical sequences from Europe but not from Switzerland: E2D (2/347), L15F (9/347) and P112S (2/347). All three mutations were detected at low mean frequencies in wastewater from Zurich (all at 3%) and Lugano (2%, 3%, and 1.7%, respectively), with E2D and L15F also detected in Geneva (3% and 5%, respectively) (Figure 3), albeit inconsistently over time (Figure S8). The most frequently observed substitution in both wastewater and clinical sequences was T12I (Europe: 123/347; Switzerland 8/34), increasing over time in wastewater from Basel, Geneva, and Laupen. Several low-frequency substitutions observed inconsistently across locations were also observed at low frequency in clinical samples. Additional low-frequent substitutions absent in clinical sequences were detected in wastewater, with K508R being the only one detected across all locations (Figure 3).

**Figure 3.**
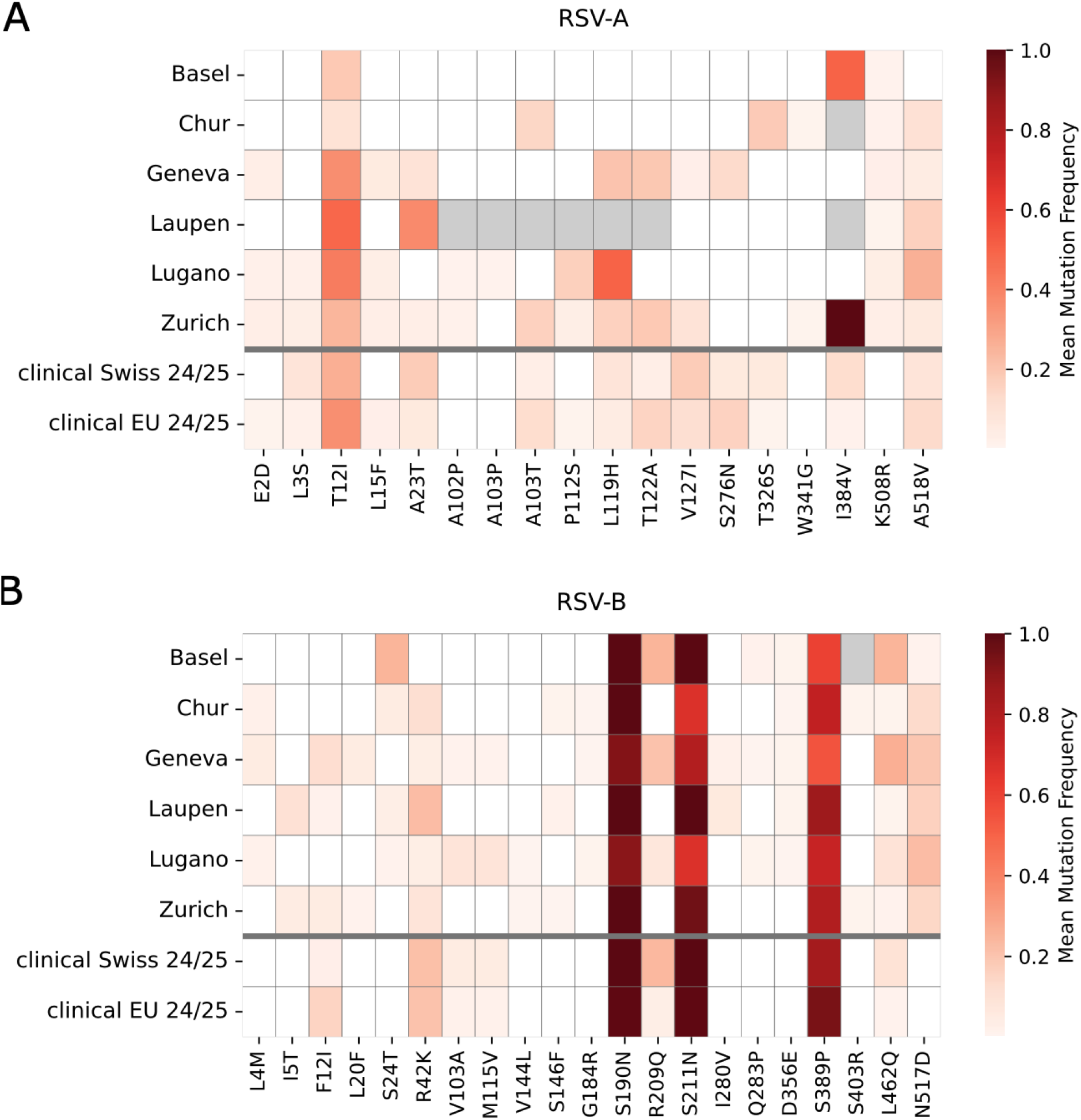
Mean frequencies over time of non-synonymous F gene mutations observed in wastewater-derived RSV-A (A) and RSV-B (B) sequences from six wastewater treatment plants across Switzerland, alongside the frequencies of these mutations in clinical Swiss and European sequences collected during the 2024-2025 RSV season. Mutations are represented as amino acid substitutions. Rows in the heatmap correspond to wastewater sampling locations or the set of clinical sequences (Swiss or European (EU)). Mutation frequencies are represented on a color scale ranging from white (lowest, 0.0) to dark red (highest, 1.0), with missing data (positions not covered in any sample at a given location throughout the 2024–2025 season) shown in grey. For wastewater data, only mutations detected in at least two wastewater samples across Switzerland, with a frequency above 0.02 and a minimum read depth of at least 10, were included in the mean frequency calculations. Mutations are reported relative to the reference EPI_ISL_412866 for RSV-A and EPI_ISL_1653999 for RSV-B.

### Detection of amino acid substitutions in the prophylaxis-targeted F gene of RSV-B

For RSV-B, the most frequent substitutions detected in both wastewater and clinical samples were S190N (Europe: 424/431, Switzerland: 42/42), S211N (Europe: 423/431, Switzerland: 42/42), and S389P (Europe: 399/431, Switzerland: 35/42), consistently detected across all locations (Figure 3 & S9). Several substitutions (F21I, R42K, V103A, M115V, R209Q, and L462Q) were observed at low frequencies in both wastewater and clinical sequences, with variable prevalences between locations. Additional low-frequency substitutions not identified in European and Swiss clinical sequences were detected in wastewater, among which N517D was the most abundant and occurred across all six locations frequently throughout the season (Figure S9).

### Differences in genetic diversity between RSV subtypes and genes

We estimated and compared the genetic diversity in wastewater samples using nucleotide diversity and mutation richness. The ratios in nucleotide diversity compare the relative accumulation of mutations in the F and G genes at the present time, while the ratios of mutation richness compare cumulative divergence since reference sequence collection. We found that RSV-A exhibited significantly higher nucleotide diversity than RSV-B across the whole genome (RSV-A/RSV-B ratio: 2.10, p.adj <0.05) and within the F gene (RSV-A/RSV-B ratio 3.14, p.adj <0.05) (Figure 4, Table S5, S6).

**Figure 4.**
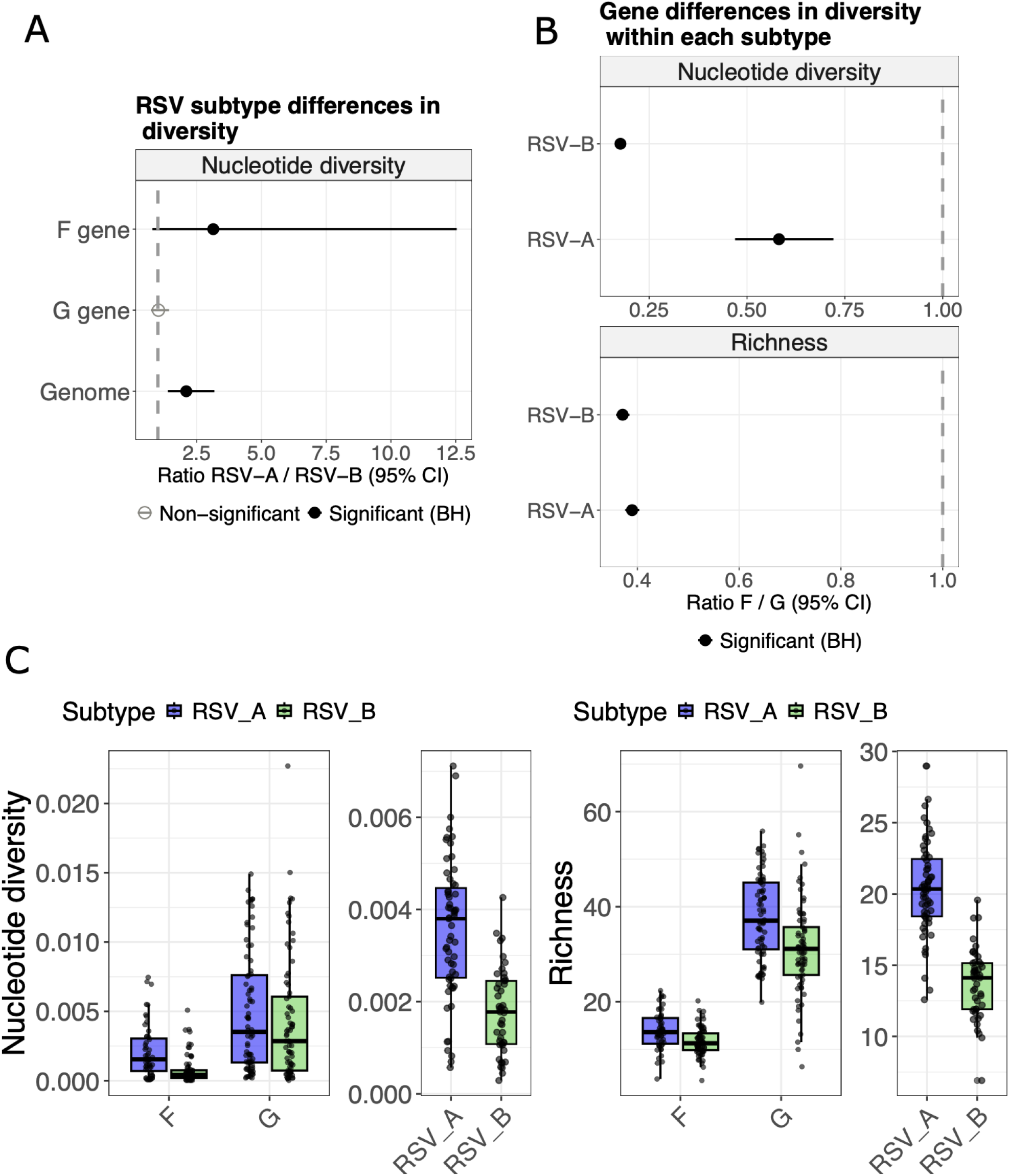
Genetic diversity comparisons between RSV subtypes and genes in wastewater-derived sequences using the nucleotide diversity metric and mutation richness. (A) Ratio between RSV-A and RSV-B diversity with 95% CI for the nucleotide diversity across the F gene, G gene, and the whole genome. (B) Ratio between F and G gene diversity with 95% CI for the nucleotide diversity and mutation richness within RSV-A and RSV-B subtypes. Black represents significant ratios different from 1 after Benjamini-Hochberg (BH) adjustment of p-values, while non-significant ratios are shown in grey. (C) Estimated nucleotide diversity and mutation richness across the F and G genes and the complete genome of RSV-A (blue) and RSV-B (green). Diversity estimates at whole-genome level required ≥50% genome coverage, and gene-level analyses required ≥50% coverage of the respective gene, both at minimum sequencing depth of ≥10 reads per position, with only positions covered ≥10 reads included in the diversity calculations.

In contrast, for the G gene, nucleotide diversity was not significantly different across subtypes. Within each subtype, diversity was consistently lower in the F gene than in the G gene across both metrics (all p.adj < 0.05). For RSV-A, the F/G diversity ratio was 0.58 for nucleotide diversity and 0.39 using richness. In contrast, for RSV-B, the F/G ratio was lower, with 0.18 for nucleotide diversity, but increased to 0.37 when using richness.

## Discussion

With the implementation of novel RSV vaccines, there is an increasing need to understand the molecular epidemiology of RSV to support monitoring vaccine efficacy. In this study, we utilize a wastewater-based RSV sequencing approach for genomic surveillance of RSV throughout the season from November 2024 until May 2025, in six locations across Switzerland, enabling tracking of RSV subtype dynamics and individual mutations with high temporal and spatial resolution.

During the 2024/2025 season, RSV-A and RSV-B co-circulated across Switzerland, with neither subtype emerging as dominant and both exhibiting similar epidemiological trajectories based on subtype-specific R_t_ estimates. While some studies describe alternating biennial dominance of RSV-A and RSV-B^37–39^, others report frequent co-circulation without clear dominance^40–42^. Several studies reported frequent predominance of RSV-A, when based on samples from patients with acute respiratory disease^41,43^. In the seasons following the COVID-19 pandemic, marked predominance of one of the subtypes coincided with increased testing^42^. Assuming similar shedding load profiles for both RSV subtypes, we also observed spatial heterogeneity, with RSV-B predominating in Geneva and RSV-A in Zurich and Lugano, consistent with previous reports on regional subtype variation^20,37^. Drivers of these temporal and geographic patterns remain unclear, as most studies are limited to short time periods, single sites or sparse clinical sampling. Our data based on regularly sampled wastewater throughout the peak season demonstrates that wastewater-based subtype surveillance can provide a more cost-effective and representative view of subtype temporal and geographic differences.

Several amino acid substitutions in the prophylaxis-targeted F protein were identified in wastewater-derived sequences. None of the substitutions currently monitored for prophylaxis resistance in Switzerland were detected in wastewater nor in clinical samples from Switzerland or Europe for both subtypes^44^. Regarding RSV-A, substitutions T12I and T122A, were detected again in 2024/2025, as in the 2023/2024 season^21^. Substitutions L20I/F, T91R, M115T, and S377N were detected in 2023/2024 but not in 2024/2025. The S377N mutation, located within the antigenic site targeted by the RSVPreF3 (AREXVY) vaccine^45^, was not identified in 2024/2025, likely because this position had sufficient sequencing coverage (≥10 reads) in only four samples over the season, although was detected in 17 out of 34 Swiss clinical RSV-A samples. Conversely, L119H and A518V were frequent in 2024/2025 after being rare or absent previously. Both have been reported in clinical sequences from a nirsevimab trial, with L119H found only in patients receiving nirsevimab^46^. Substitution K508R, which has been reported before^46^, was detected in wastewater, but not in clinical sequences.

For RSV-B, substitutions R42K, S190N, S211N, R209Q, and S389P were frequently detected, similar to the 2022-2023 season^21^. S211N and R209Q, signature mutations of, respectively, the B.D.E.1 and B.D/B.D.1 sublineages, are both located on the antigenic site of Nirsevimab^45^ but are not linked to resistance, except if S211N co-occurs with K65Q^47^ which was not detected. Additional low-frequency substitutions were identified in 2024/2025, whereas I431L and T555A were no longer detected in 2024/2025. Newly identified L462Q and N517D appeared consistently across locations (Figure S9). Notably, N517D has not been reported in Swiss or European clinical sequences from 2024/2025 and neither in previously reported sequences^46^. Although wastewater sequencing data can be noisy, mutations occurring consistently over time and across locations increase the confidence in their detection. This underscores the ability of wastewater sequencing to identify mutations within locally circulating sublineages that affect F protein antigenic sites, which may go undetected in clinical genomic surveillance.

Wastewater sequencing data also allows for assessing the genomic diversity of samples and hence indirectly of the viral strains infecting the host population in a wastewater-treatment plant catchment area. We found greater whole-genome and F-gene diversity in RSV-A than RSV-B, consistent with previous clinical reports^48,49^. In contrast, G gene diversity, typically the most variable region^50^, did not differ significantly between subtypes, diverging from earlier studies reporting higher G-gene diversity in either RSV-A^48^ or RSV-B^39^. These discrepancies indicate that observed diversity patterns depend strongly on the underlying dataset and the methods used. However, there may also be an impact of differences in evolutionary processes between subtypes, including different mutation rates and selective pressures, though underlying factors remain to be elucidated. Different diversity metrics were used, with nucleotide diversity (π) measuring average pairwise distances between sequences and reflecting within-population variation at the sampling time. Mutation richness instead quantifies the total number of distinct mutations relative to the reference, representing cumulative divergence since the reference sequence was collected. For RSV-A, the F/G diversity ratio was lower using the richness metric compared to π, whereas for RSV-B, the opposite pattern was observed. In addition, while the richness-based ratio was similar for both subtypes, the π-based ratio for RSV-A was higher than for RSV-B. This suggests that while both subtypes accumulated similar proportions of mutations in F vs G over evolutionary time, due to subtype-specific evolutionary dynamics^49^, the current circulating RSV-B population shows less within-population variation in the F gene relative to G than RSV-A.

A major limitation of our study was the low median genome coverage across samples, with 0.16 for RSV-A and 0.17 RSV-B, increasing to 0.34 for RSV-A and 0.31 for RSV-B, for samples above the dPCR subtyping assay’s detection limit (Figure S7). Coverage correlated strongly with subtype concentration, indicating that co-circulation, and thus lower per-subtype concentrations, were the main reason for reduced coverage compared with our previous work^21^. Unlike our previous study, in which samples with high concentrations were selected for sequencing, samples in this study were selected randomly, yielding estimates unbiased by sampling strategy. However, the lower concentrations in the randomly selected samples likely contribute to the lower overall coverage, which led to dropouts and missed mutations, potentially leaving clinically relevant mutations undetected. Consistent with prior observations, wastewater sequencing performs best with elevated concentrations, suggesting the use of a threshold when selecting samples for sequencing, such as those based on concentrations measured by dPCR. Despite the low coverage, we were still successful in tracking individual mutations on clinically relevant sites as well as identifying differences in genetic diversity across subtypes and genes. In addition, the number of available clinical sequences collected in Switzerland was low for both RSV-A and RSV-B (34 and 42, respectively, (Figure S4)), which may have biased the frequency of mutations reported in clinical Swiss data.

Extending the presented wastewater-based genomic RSV surveillance approach into future RSV seasons with expanding geographic scope will enable continuous tracking of viral dynamics, which is particularly important given the widespread introduction of new immunoprophylaxis products. Going forward, we envision wastewater genomic surveillance as a core complement to clinical sequencing, offering cost-effective population-level insights into RSV subtype circulation and evolutionary trends.

## Data availability

Digital PCR data of viral loads in wastewater is available for download from wise.ethz.ch. Wastewater sequencing data used in this study is available on ENA under project number PRJEB85524. The data analysis code and scripts used for data visualization are available on Github under https://github.com/cbg-ethz/RSV-wastewater-2024-2025.

## Acknowledgments

We would like to thank all members of the Wastewater-based Infectious disease Surveillance (WISE) consortium.

## Funding

This study was funded by the Swiss National Science Foundation (Sinergia grant CRSII5_205933). Funding for sample collection and processing was provided by the Swiss Federal Office of Public Health.

## Competing interests

None

## Notes

### Competing Interest Statement

The authors have declared no competing interest.

