## Supplementary material for "Tracking Respiratory Syncytial Virus dynamics in wastewater during the 2024-2025 season in Switzerland"

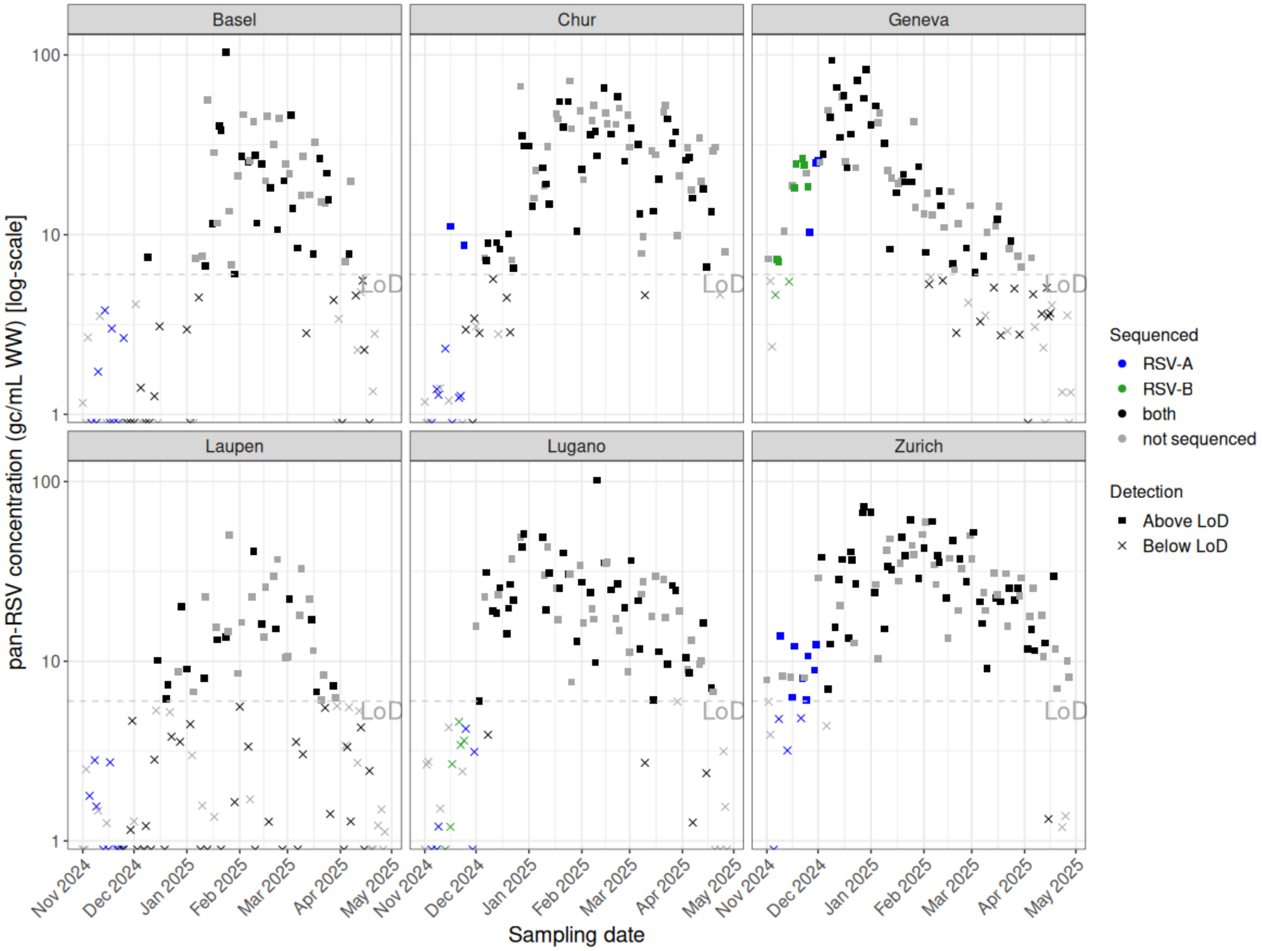


**Figure S1. Selection of 328 wastewater samples for RSV sequencing, from six locations across Switzerland during the 2024-2025 RSV season.** Pan-RSV concentrations were measured as part of the Swiss wastewater-based respiratory virus monitoring program. Initially, only the predominant subtype was sequenced, shown in blue (RSV-A) or green (RSV-B), as determined by a subtype specific digital PCR assay. After November 19, 2024, both subtypes were sequenced in all samples (black). The pan-RSV digital PCR assay had a limit of detection (LoD) of six genome copies per mL of wastewater, corresponding to at least three positive partitions, shown as a dashed grey line.

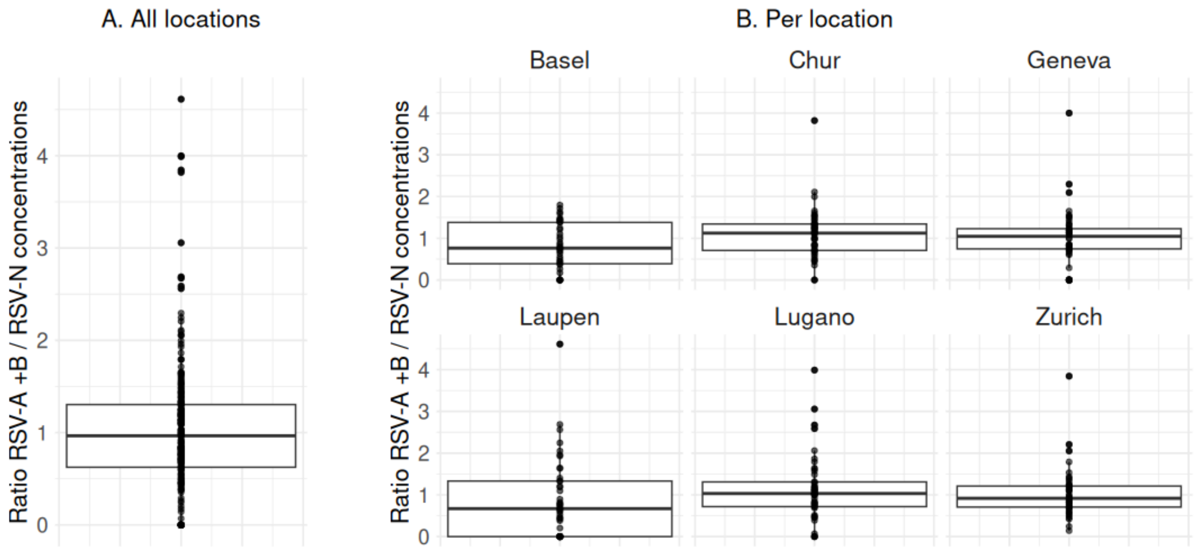


**Figure S2. Ratio between concentrations measured with the pan-RSV dPCR assay (RSV-N) and the summed concentrations for RSV-A and RSV-B, measured with the subtype-specific dPCR assay.** (A) Ratios across all locations combined. (B) Ratios separated by individual locations.

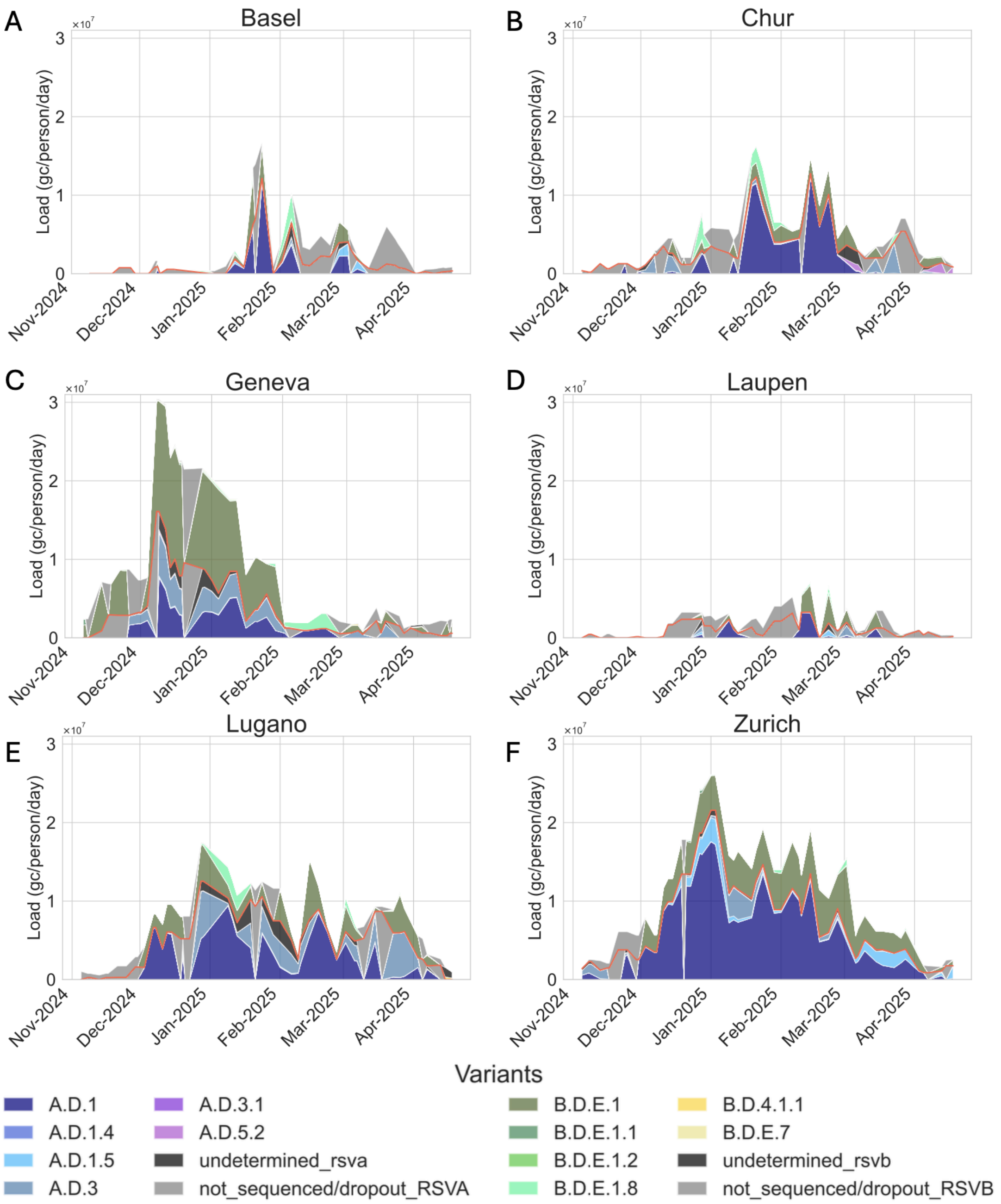


**Figure S3. Temporal dynamics of circulating RSV-A and RSV-B lineages in wastewater from six locations across Switzerland during the 2024-2025 RSV season.** (A-F) Estimated abundances of RSV-A (blue-red shades) and RSV-B (green-yellow shades) lineages, calculated by scaling lineage proportions. Proportions were inferred from signature mutation frequencies (Pathoplexus accession date: 15 Oct 2025) using Lollipop version 0.5.0[1], as previously described[2]. , and scaled to the 7-day median flow-normalized RSV subtype concentrations measured with a subtype-specific digital PCR assay. Lineages were considered for deconvolution only if present in the publicly available Swiss clinical sequences (Figure S4). Samples not sequenced for a given subtype or dropouts with zero coverage, are shown in light grey. RSV-A and RSV-B proportions are separated by a red solid line.

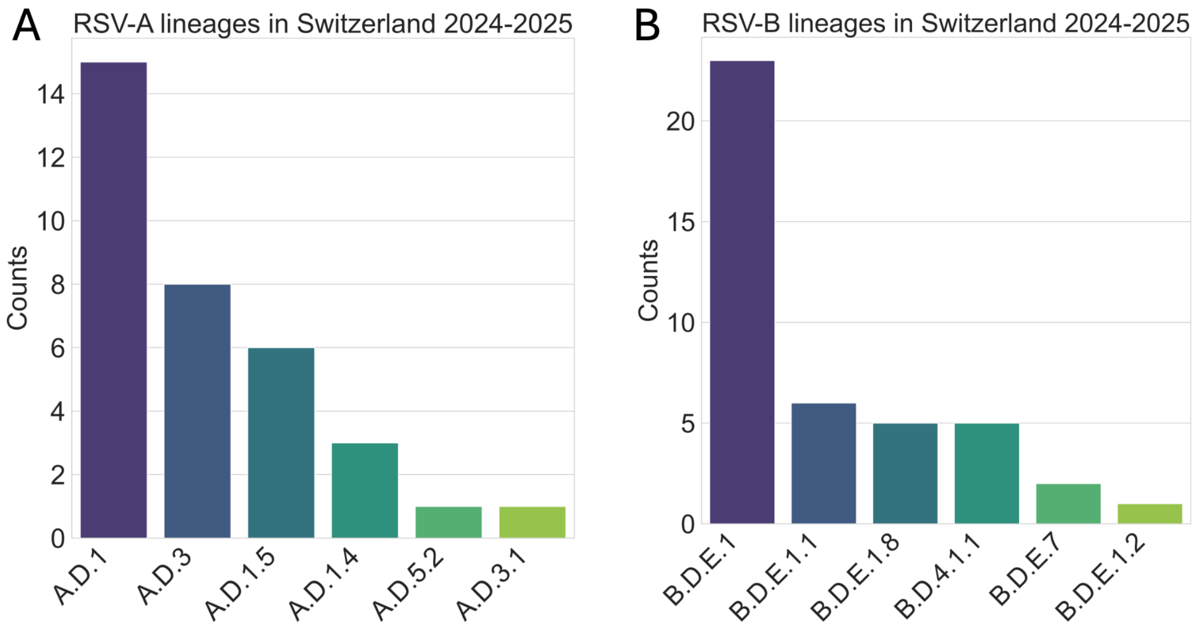


**Figure S4. Number of clinical sequences assigned to (A) RSV-A and (B) RSV-B lineages in Switzerland, collected between 1 October 2024 and 30 April 2025.** In total, 34 RSV-A and 42 RSV-B sequences were available in the Pathoplexus database (accession date 12 Oct 2025)**.**

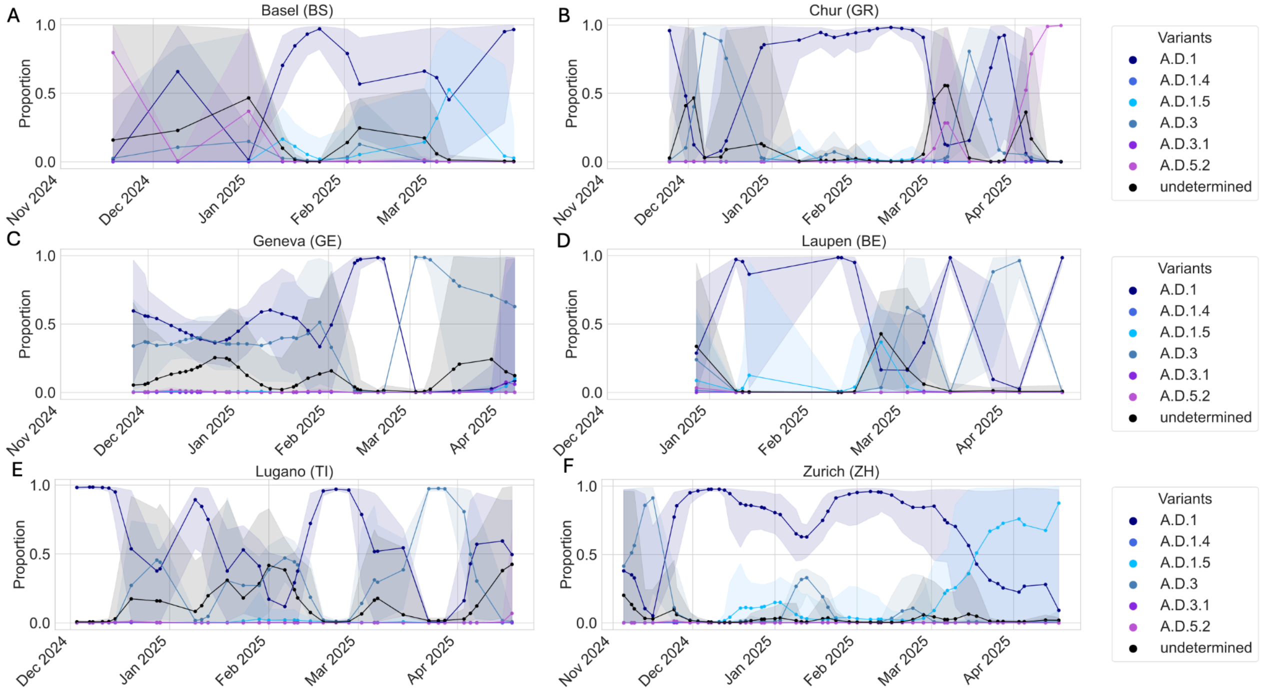


**Figure S5. Estimating relative abundances of RSV-A lineages over time from the signature mutation frequencies.** (A-F) Estimated proportions of RSV-A lineages in wastewater collected from six locations across Switzerland during the 2024-2025 RSV season. The shaded bands around the lines represent the 95% confidence intervals around the proportion estimates, based on bootstrapping.

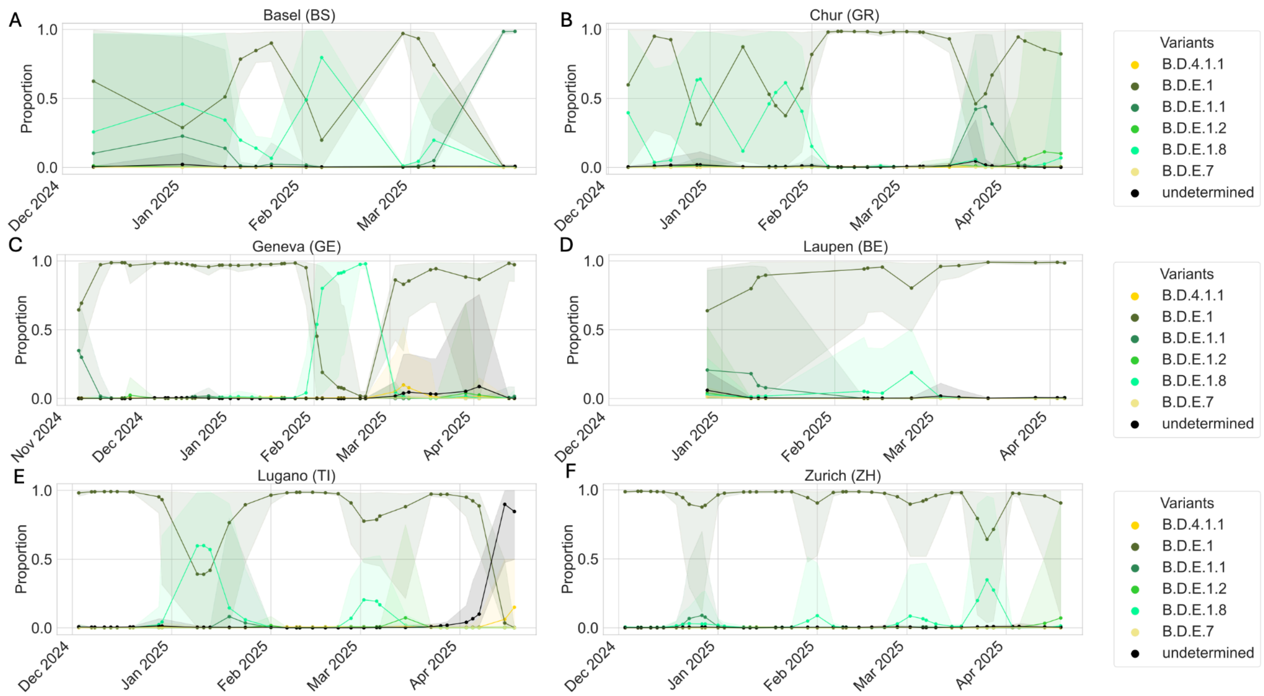


**Figure S6. Estimating relative abundances of RSV-B lineages over time from the signature mutation frequencies.** (A-F) Estimated proportions of RSV-B lineages in wastewater collected from six locations across Switzerland during the 2024-2025 RSV season. The shaded bands around the lines represent the 95% confidence intervals around the proportion estimates, based on bootstrapping.

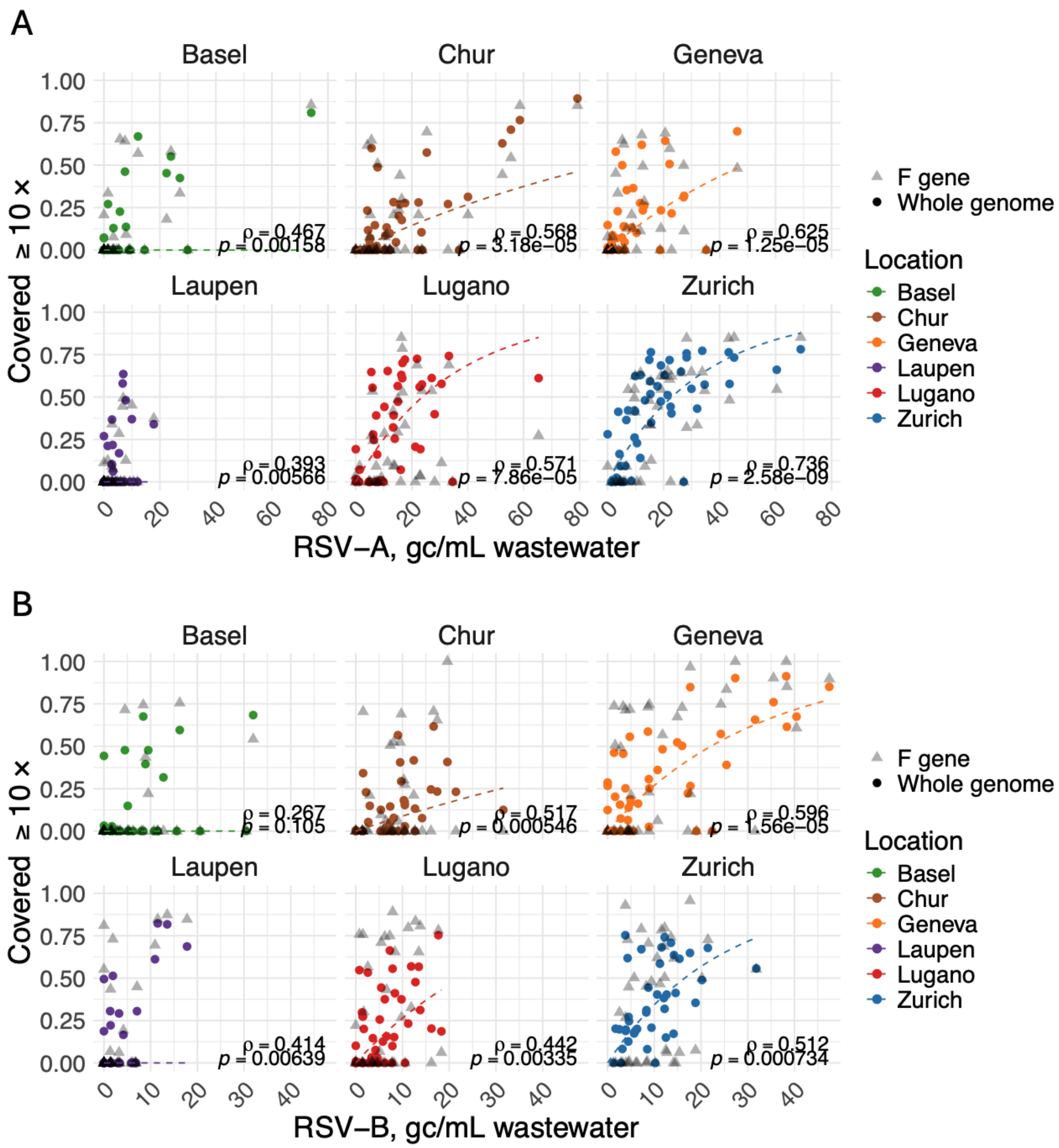


**Figure S7. Proportion of the reference genome covered at a read depth of at least 10 for (A) RSV-A and (B) RSV-B versus the subtype concentrations (in genome copies per mL wastewater) measured with the subtype-specific dPCR assay.** Triangles correspond to the proportion of F-gene covered, and circles the covered proportion of the full genome. Only samples with known subtype-specific concentrations were included. The dashed lines represent the fitted exponential saturation curves $E[cov] = 1 - e^{-conc * k}$, based on the whole genome coverage, and are added to aid visualization. The curves were modeled using robust linear regression of the form *−log(1 − cov) ~ conc*, implemented with the R package robustbase (version 0.99-2). Computed nonparametric Spearman correlation coefficients and p-values are provided at the bottom right.

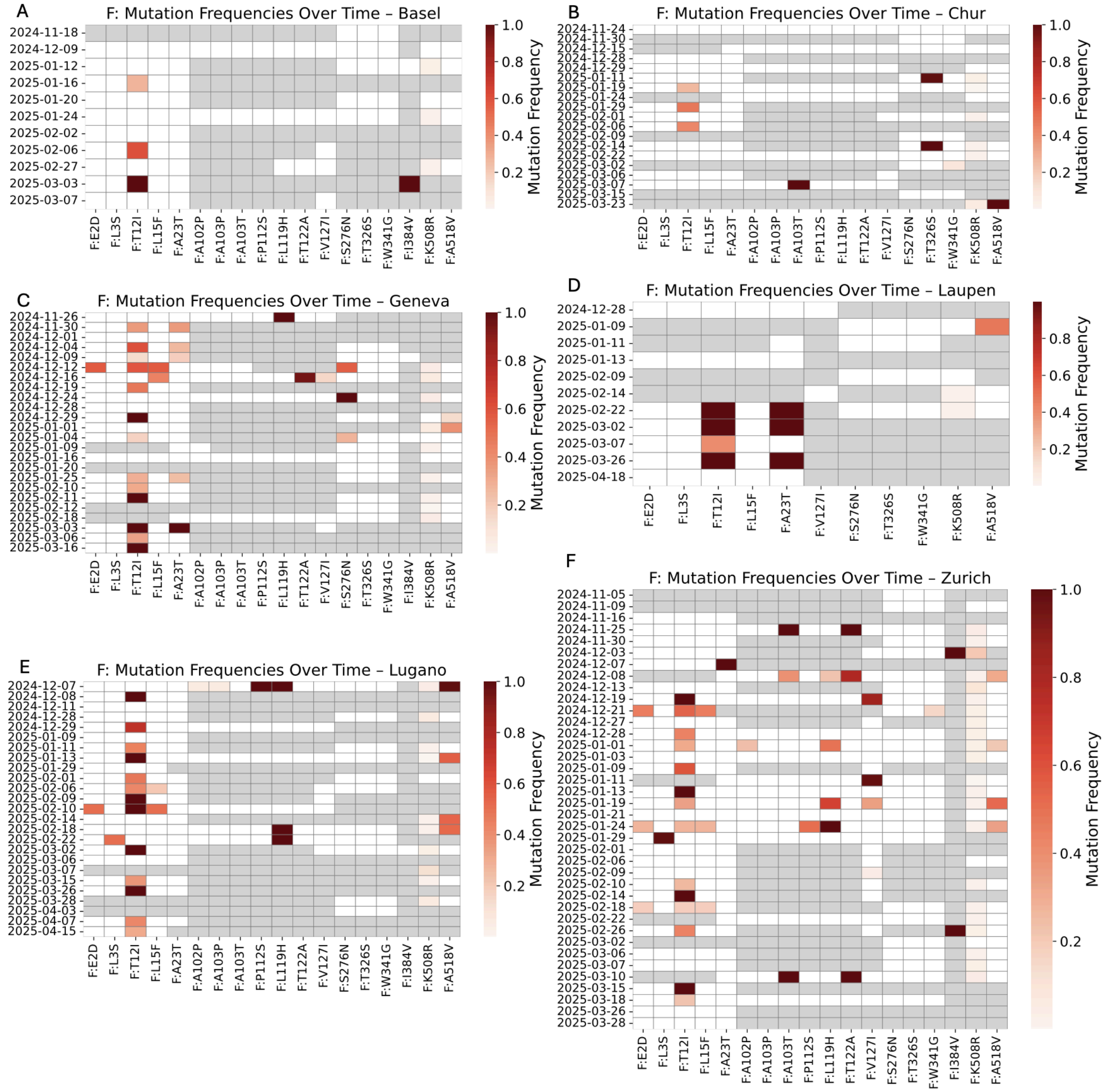


**Figure S8. Amino acid substitution frequencies in the RSV-A fusion (F) protein, detected in wastewater samples from six cities across Switzerland during the 2024-2025 RSV season.** Rows refer to the sampling date, while columns represent the substitution. Frequencies are encoded from white (lowest, 0.0) to dark red (highest, 1.0), while dropouts (read depth below 10 per position) are shown in grey. Substitutions are only visualized when detected in at least two samples with a frequency above 0.02 and a read depth of at least 10. Mutations are reported relative to the reference EPI_ISL_412866.

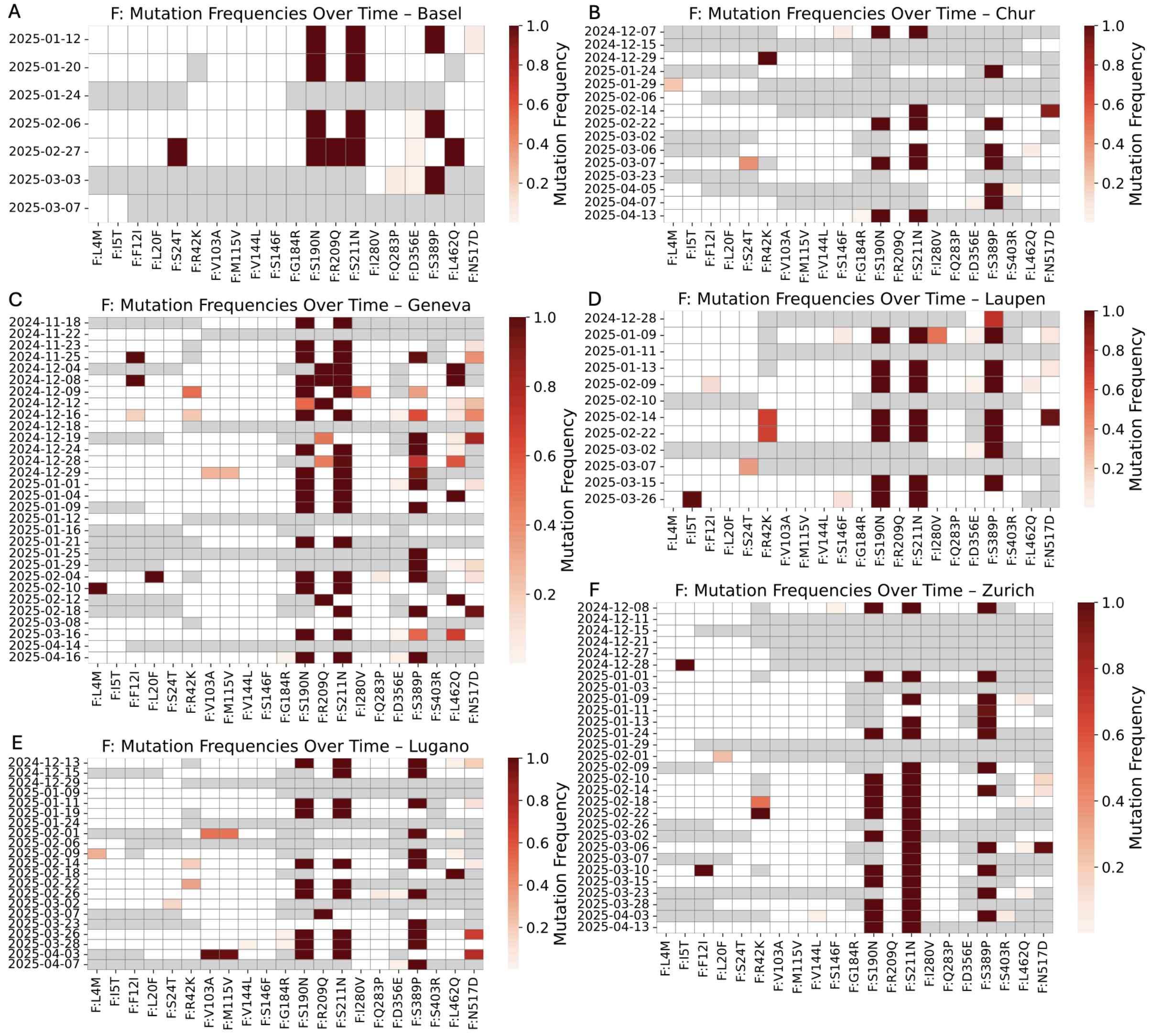


**Figure S9. Amino acid substitution frequencies in the RSV-B fusion (F) protein, detected in wastewater samples from six cities across Switzerland during the 2024-2025 RSV season.** Rows refer to the sampling date, while columns represent the substitution. Frequencies are encoded from white (lowest, 0.0) to dark red (highest, 1.0), while dropouts (read depth below 10 per position) are shown in grey. Substitutions are only visualized when detected in at least two samples with a frequency above 0.02 and a read depth of at least 10. Mutations are reported relative to the reference EPI_ISL_1653999.

**Table S1. Catchment-specific calibrated average detectable load shed over the course of infection (gc = gene copies), based on Sentinella reporting and participatory surveillance data in Switzerland.**

| Location | Calibrated average detectable load shed |
|---|---|
| Basel | $8.90 \times 10^9$ gc |
| Chur | $14.54 \times 10^9$ gc |
| Geneva | $13.39 \times 10^9$ gc |
| Laupen | $7.00 \times 10^9$ gc |
| Lugano | $17.70 \times 10^9$ gc |
| Zurich | $24.07 \times 10^9$ gc |

**Table S2. Generalized linear mixed-effects models (GLMMs) fitted to compare genetic diversity metrics between RSV subtypes and genes.**

| Comparison | Diversity metrics (response variables) | Fixed effects | Random effect | Family (link) |
|---|---|---|---|---|
| RSV-A vs RSV-B, whole genome | Nucleotide diversity | Subtype, log(mean coverage), log(mean coverage)^2 | location | Gamma (log) |
| RSV-A vs RSV-B, F gene | Nucleotide diversity | Subtype, log(mean coverage), log(mean coverage)^2 | location | Gamma (log) |
| RSV-A vs RSV-B, G gene | Nucleotide diversity | Subtype, log(mean coverage), log(mean coverage)^2 | location | Gamma (log) |
| F vs G, RSV-A | Nucleotide diversity, richness | Gene (F/G), log(mean coverage), log(mean coverage)^2 | location | Gamma (log) |
| F vs G, RSV-B | Nucleotide diversity, richness | Gene (F/G), log(mean coverage), log(mean coverage)^2 | location | Gamma (log) |

**Table S3. Estimated marginal means with 95% confidence intervals for each diversity metric, derived from GLMMs.** All means are back-transformed to the original scale from the log scale.

*Marginal means are model-estimated and depend on the data subset used for each comparison context. Estimates for the same biological group may slightly differ across comparison types due to differences in model structure and the data used for parameter estimation.

| comparison_type | subtype | gene | group | metric | response | asymp.LCL | asymp.UCL |
|---|---|---|---|---|---|---|---|
| Subtype, whole genome | RSV_A | | Whole genome | Nucleotide diversity | 0.0033 | 0.00272 | 0.004 |
| Subtype, whole genome | RSV_B | | Whole genome | Nucleotide diversity | 0.00158 | 0.00125 | 0.00198 |
| Subtype, gene level | RSV_A | | F gene | Nucleotide diversity | 0.00224 | 0.00155 | 0.00323 |
| Subtype, gene level | RSV_B | | F gene | Nucleotide diversity | 0.00071 | 0.00051 | 0.001 |
| Subtype, gene level | RSV_A | | G gene | Nucleotide diversity | 0.00394 | 0.00307 | 0.00506 |
| Subtype, gene level | RSV_B | | G gene | Nucleotide diversity | 0.00386 | 0.00298 | 0.00499 |
| Gene, within subtype | | F | RSV-A | Nucleotide diversity | 0.00213 | 0.00159 | 0.00287 |
| Gene, within subtype | | G | RSV-A | Nucleotide diversity | 0.00367 | 0.00278 | 0.00484 |
| Gene, within subtype | | F | RSV-B | Nucleotide diversity | 0.0007 | 0.00051 | 0.00097 |
| Gene, within subtype | | G | RSV-B | Nucleotide diversity | 0.00398 | 0.00285 | 0.00556 |
| Gene, within subtype | | F | RSV-A | Richness | 14.47136 | 13.4486 | 15.5719 |
| Gene, within subtype | | G | RSV-A | Richness | 37.13717 | 34.76182 | 39.67484 |
| Gene, within subtype | | F | RSV-B | Richness | 11.53857 | 10.64608 | 12.50588 |
| Gene, within subtype | | G | RSV-B | Richness | 31.0926 | 28.54862 | 33.86327 |

**Table S4. Subtype proportions measured with dPCR across six Swiss cities during the 2024-2025 RSV season.**

| Location | Subtype | IQ1 | Median | IQ3 |
|---|---|---|---|---|
| Basel | RSV-A | 0.28 | 0.49 | 0.63 |
| | RSV-B | 0.37 | 0.51 | 0.72 |
| Chur | RSV-A | 0.50 | 0.66 | 0.82 |
| | RSV-B | 0.18 | 0.34 | 0.50 |
| Geneva | RSV-A | 0.26 | 0.36 | 0.52 |
| | RSV-B | 0.48 | 0.64 | 0.74 |
| Laupen | RSV-A | 0.46 | 0.58 | 0.90 |
| | RSV-B | 0.10 | 0.42 | 0.54 |
| Lugano | RSV-A | 0.53 | 0.70 | 0.85 |
| | RSV-B | 0.15 | 0.30 | 0.47 |
| Zurich | RSV-A | 0.53 | 0.70 | 0.77 |
| | RSV-B | 0.23 | 0.30 | 0.47 |

**Table S5. Descriptive statistics for diversity metrics – nucleotide diversity and richness – derived from wastewater sequencing data and summarized as median (Q1 - Q3) for RSV-A and RSV-B subtypes at the whole-genome level and across F, G genes separately.**

| genome_region | subtype | n |
|---|---|---|
| F gene | RSV-A | 51 |
| F gene | RSV-B | 69 |
| G gene | RSV-A | 76 |
| G gene | RSV-B | 73 |
| Whole genome | RSV-A | 59 |
| Whole genome | RSV-B | 45 |
| **genome_region** | **subtype** | **Nucleotide_diversity** |
| F gene | RSV-A | 0.0015 (7e-04–0.003) |
| F gene | RSV-B | 4e-04 (2e-04–8e-04) |
| G gene | RSV-A | 0.0035 (0.0013–0.0076) |
| G gene | RSV-B | 0.0029 (7e-04–0.0061) |
| Whole genome | RSV-A | 0.0038 (0.0025–0.0045) |
| Whole genome | RSV-B | 0.0018 (0.0011–0.0024) |
| **genome_region** | **subtype** | **Richness** |
| F gene | RSV-A | 13.624 (11.1802–16.5856) |
| F gene | RSV-B | 11.2782 (9.8551–13.3753) |
| G gene | RSV-A | 37.0551 (31.041–45.0715) |
| G gene | RSV-B | 31.1355 (25.641–35.7143) |
| Whole genome | RSV-A | 20.3541 (18.4329–22.4529) |
| Whole genome | RSV-B | 14.1156 (11.9168–15.1457) |

**Table S6. Pairwise contrasts from GLMMs comparing diversity metrics between RSV subtypes (RSV-A vs RSV-B) or between genes (F vs G).** Ratios are back transformed from the log scale; values greater than one indicate higher diversity in the first group (RSV-A or F gene, respectively). P-values were adjusted for multiple comparisons using the Benjamini-Hochberg procedure applied separately within each comparison type (subtype comparisons and gene comparisons). Asterisk (*) indicates adjusted p-value < 0.05.

| fixed_effect | genomic_level | metric | compared | ratio | SE | z.ratio | p.value | p.adjusted | significant |
|---|---|---|---|---|---|---|---|---|---|
| Subtype | Whole genome | Nucleotide diversity | RSV_A / RSV_B | 2.096504 | 0.212972 | 7.28725 | 3.16E-13 | 3.163E-13 | * |
| Subtype | F gene | Nucleotide diversity | RSV_A / RSV_B | 3.137913 | 0.706866 | 5.076468 | 3.85E-07 | 7.69E-07 | * |
| Subtype | G gene | Nucleotide diversity | RSV_A / RSV_B | 1.020572 | 0.172965 | 0.120154 | 0.904361 | 0.9043609 | |
| Gene | RSV-A | Nucleotide diversity | F / G | 0.581653 | 0.109341 | -2.88262 | 0.003944 | 0.0039438 | * |
| Gene | RSV-B | Nucleotide diversity | F / G | 0.176471 | 0.032658 | -9.37311 | 7.04E-21 | 9.39E-21 | * |
| Gene | RSV-A | Richness | F / G | 0.389673 | 0.018207 | -20.1705 | 1.78E-90 | 3.556E-90 | * |
| Gene | RSV-B | Richness | F / G | 0.371103 | 0.017881 | -20.5728 | 4.81E-94 | 1.924E-93 | * |
